# Knowledge and attitude towards COVID-19 in Bangladesh: Population-level estimation and a comparison of data obtained by phone and online survey methods

**DOI:** 10.1101/2020.05.26.20104497

**Authors:** Anwarul Karim, Mastura Akter, AHM Thafikul Mazid, Orindom Shing Pulock, Tasmiah Tahera Aziz, Samira Hayee, Nowrin Tamanna, GS Chuwdhury, Afsana Haque, Farhana Yeasmin, Mashkura Akter Mitu, Farjana Yeasmin, Humayun Rashid, Ashish Kumar Kuri, Arni Das, Koushik Majumder, Dipen Barua, Md Mahabubur Rahaman, Sanjida Akter, Nashid Niaz Munia, Jabin Sultana, Faeeqa Usaila, Sabrina Sifat, Nishat Anjum Nourin, Md Forhad Uddin, Mrinmoy Bhowmik, Tanvir Ahammed, Nabil Sharik, Quddus Mehnaz, Md Nur Hossain Bhuiyan, Tahmina Banu

## Abstract

Peoples’ adherence to the guidelines and measures suggested in fighting the ongoing COVID-19 pandemic is partly determined by the Knowledge, Attitude, and Practices (KAP) of the population. In this cross-sectional study, we primarily addressed two key issues. First, we tried to determine whether there is a significant difference in the estimated COVID-19 knowledge level from the online and phone survey methods. Second, we tried to quantify the knowledge and attitude of COVID-19 in Bangladeshi adult population. Data were collected through phone calls (April 14-23, 2020) and online survey (April 18-19, 2020) in Bangladesh. The questionnaire had 20 knowledge questions with each correct response getting one point and incorrect/don’t know response getting no point (maximum total knowledge score 20). Participants scoring >17 were categorized as having good knowledge. The percentages of good knowledge holders were 57.6%, 75.1%, and 95.8% in the phone (n=1426), online non-medical (n=1097), and online medical participants (n=382), respectively. Comparison between phone and online survey showed that, overall, online survey might overestimate knowledge level than that of phone survey, although there was no difference for elderly, poor, and rural people. Male gender, higher education, living in town/urban areas, good financial condition, and use of internet were positively associated with good knowledge. However, higher knowledge was associated with having less confidence in the final control of COVID-19. Our adult population-level estimates showed that only 32.6% (95% CI 30.1-35.2%) had good knowledge. This study provides crucial information that could be useful for the researchers and policymakers to develop effective strategies.

## 1. Background

Coronaviruses (CoVs) are a large family of enveloped, positive-sense RNA viruses that are important pathogens of humans and other mammals [1]. In the past two decades, two highly pathogenic human CoVs, namely SARS-CoV and MERS-CoV, have emerged in 2003 and 2012 respectively [2]. Recently, a third new type of CoV, which is even more infectious, is spreading across the world in an unprecedented manner. This novel coronavirus was discovered to be the cause of unexplained pneumonia-like cases in Wuhan, Hubei, China in December, 2019 with the majority of initial patients had been exposed to Huanan seafood market [3]. This coronavirus, which was provisionally named as 2019-nCoV, has been renamed as Severe Acute Respiratory Syndrome Coronavirus 2 (SARS-CoV-2). The disease caused by SARS-CoV-2 has been named as coronavirus disease 2019 (COVID-19) [4]. It was first reported to the World Health Organization (WHO) on December 31, 2019. As of April 30, globally confirmed cases rose to 3 090 445 (with 217 769 deaths) from 212 countries/territories/areas [5]. Bangladesh announced the first three cases of COVID-19 on March 8, 2020. Just by April 30, the total number of confirmed cases raised to 7667 (with 168 deaths) [6].

To reduce the spread of COVID-19 and chance of being infected, WHO advises public to adopt some simple precautions of maintaining hygiene and social distancing [7]. Authorities from different countries around the world are providing additional advice and adopting further measures e.g., complete or partial shutdown of different areas, restricting transport facilities, travel bans, transitioning educational and business activities to online, restricting access to public places. An important determinant of peoples’ adherence to these guidelines and measures could be the Knowledge, Attitudes, and Practices (KAP) towards COVID-19 of the respective population [8-10]. Furthermore, information about KAP of the general population may guide the policymakers and researchers tremendously to develop effective strategies to face this kind of rapidly evolving crisis.

Some COVID-19 KAP survey reports from different countries [11-20], including Bangladesh [21, 22], are available already. However, most of these surveys were conducted through online questionnaire as it was not so feasible to do it otherwise during this pandemic disruption. Online surveys probably have limited value when estimating population-level KAP in countries like Bangladesh, where only 15% of the population has access to internet compared to the world average of 50% (World Bank – 2017) [23]. Therefore, KAP of the general Bangladeshi population towards COVID-19 is largely unknown and it is imperative to conduct further studies to understand more about it so that strategies can be developed or adapted accordingly. In this study, we surveyed adults from Bangladesh through phone call interviews and an online questionnaire. This allowed us to investigate whether one method tends to overestimate knowledge level over another. The survey with phone calls enabled us to access people from diverse sociodemographic backgrounds, thus allowing us to adjust the phone survey data to the population-level sociodemographic characteristics to estimate COVID-19 knowledge status of the adult population. To the best of our knowledge, this study is the largest in Bangladesh conducted so far to reveal the knowledge and attitudes of its adult population towards COVID-19 during its rapid rise period and the only study to do a comparative evaluation of phone and online survey methods in estimating COVID-19 knowledge.

## 2. Methods

Ethical clearance was obtained from the ethical review committee of Chittagong Medical College, Chattogram, Bangladesh. A cross-sectional survey was conducted through phone calls from April 14-23, 2020. In addition, an online survey was conducted from April 18-19, 2020. As it was difficult to predict the number of participants we would be able to recruit during this disruption, we had no predetermined target sample size.

### 2.1. Phone survey

Due to the rapid spread of COVID-19 and significant disruption (termed as “lockdown” by many, but not officially) of different areas of Bangladesh during our study period, it was not feasible to conduct a community-based national sampling survey. Therefore, we had to rely on the authors’ network to recruit participants. For better representation of the diverse population and to cover all the administrative divisions of Bangladesh, we first formed a research group with members studied/studying/worked/working in various universities/institutes throughout the country. Members were trained to conduct the phone survey with same instructional videos and materials so that everyone could follow a similar approach while conducting the survey. Inclusion criteria were: a) persons giving consent to participate, and b) at least 18 years old. Exclusion criteria were: a) persons not giving consent, b) persons who could not communicate well, c) researcher’s family members, d) researcher’s relatives/friends/colleagues with whom the researcher previously discussed about COVID-19 to make them aware, e) other family members/friends/colleagues living together with already included participant, and f) doctors/nurses/medical students and family members living with them. The researchers used their acquaintances, acquaintances of family members or friends or colleagues, and acquaintances of acquaintances to recruit a wide range of participants of different age groups, geographic locations, financial conditions, and educational backgrounds. When we requested our acquaintances to suggest some of their acquaintances to be included in our study, they were requested not to disclose the survey questions asked from them during their participation in our survey. Conversations with the participants were made through phone calls. At the beginning, we obtained informed verbal consent. After that, we asked whether they had heard about the recent outbreak of COVID-19 or SARS-CoV-2. If they answered “yes”, then we proceeded with the questionnaire which took on average 10-12 minutes to complete. If the participants appeared to be not communicating well enough or not understanding the questions reasonably, we excluded them from the survey. After the end of the interview, they were informed briefly about the important information regarding COVID-19 and the correct answers to the questions asked.

### 2.2. Online Survey

The online survey was done with the same questionnaire with additional attitude questions that were not covered in the phone survey (to minimize phone call duration). The questionnaire was created on Google Forms and distributed through social media. The online link was posted and reposted on the Facebook timeline of the researchers, different Facebook groups, sent through friends or acquaintances through Facebook messenger or WhatsApp. Besides, the researchers requested their acquaintances to post/share the survey link on their Facebook timeline or social media apps. The link was accompanied by a summary of the study to allow the participants to make an informed decision. We requested Bangladeshi citizens who understood the nature of the study, was willing to participate, and was not surveyed by the phone call to attempt the questionnaire. At the beginning of the survey, they were asked whether they had heard about the recent outbreak of COVID-19 or SARS-CoV-2. If they answered “yes”, then the questions of the questionnaire appeared section by section and at the end of the survey, they were provided with important information regarding COVID-19 and referred to the website of WHO and Institute of Epidemiology, Disease Control and Research (IEDCR) of Bangladesh for further information. If they answered “no”, they were directly forwarded to the last information page.

### 2.3. Measures

The sociodemographic data collected in both phone and online surveys included the age in years (≥18 to <35, ≥35 to <55, and ≥55), gender (male, female, other), education level (≤ grade 5, grade 6 to 12, > grade 12 to Bachelor, > Bachelor to Master/above), lived mostly in the last four months (rural, urban/town areas, other), financial condition (poor, middle-class, rich), and administrative division where the participant lived most of the life. Information about occupation (doctor/nurse/medical student, others) was asked in online survey. This auxiliary information allowed us to stratify the online survey data to “online non-medical” and “online medical” participants. Because participants of <18 years old could not be restricted from attempting the questionnaire, we added one more age category (<18 years) in the online survey. This enabled us to exclude them from data analysis as our targets were the participants of ≥18 years old.

The questionnaire is given in Table 1. It was developed based on the information provided by the WHO, IEDCR, and Centers for Disease Control and Prevention (CDC) of USA to the general public, latest scientific evidence from literature, and previously published similar study [10]. Both English and Bangla versions of the questionnaire were agreed by a panel comprising of one clinical professor, two epidemiologists, five physicians, two medical students, and four non-medical researchers. It was tested by a pilot survey from 20 participants through phone call to ensure that all the questions were clearly understood. It is worth mentioning that there are several dialects of Bangla language spoken throughout the country. During the phone survey, sometimes researchers had to conduct survey using the local dialect.

**Table 1.** Questionnaire used in the present study

|  |  |
| --- | --- |
|  | **Section-1: Source of information regarding COVID-19 or SARS-CoV-2 (can choose more than one)** |
|  | a. Television b. Printed newspapers c. Internet (including social media e.g., Facebook)<br>d. From other people e. Other means |
|  | **Knowledge and attitude questions** |
|  | **Section-2A: Questions related to the knowledge of the disease (KD)** |
| KD01 | Some patients may have no symptoms (true) |
| KD02 | Fever is a common symptom of this disease (true) |
| KD03 | Cough is a common symptom of this disease (true) |
| KD04 | Patients with this disease can suffer from shortness of breath or difficulty in breathing (true) |
| KD05 | Some patients with this disease can have tiredness, sore throat, headaches, body aches (true) |
| KD06 | Some patients with this disease can have diarrhea (true) |
| KD07 | This disease can be completely prevented by taking antibiotics or other medicine (false) |
| KD08 | It can take up to 14 days after being infected to develop symptoms (true) |
| KD09 | The disease can spread from an infected person to others even before symptoms develop (true) |
| KD10 | This virus can spread from an infected person through talking or breathing (true) |
| KD11 | This virus can spread from an infected person through coughing (true) |
| KD12 | People can get infected by touching objects or surfaces contaminated with this virus, and then touching their eyes, nose or mouth without cleaning hands (true) |
| KD13 | All the patients with this disease die (false) |
| KD14 | Older persons and persons with heart disease, lung disease, diabetes are at high risk of developing severe COVID-19 disease (true) |
|  | **Section-2B: Questions related to the knowledge of the preventive practices (KP)** |
| KP01 | Younger persons do not need to follow any precautionary measures to prevent the disease (false) |
| KP02 | Washing hands frequently and thoroughly with soap or alcohol-based hand sanitizers reduces the risk of being infected by this virus (true) |
| KP03 | To reduce the risk of being infected by this virus, one should avoid touching eyes, nose, mouth with uncleaned hands (true) |
| KP04 | To reduce the spread of this virus, one should cover his mouth and nose with bent-elbow or tissue paper or handkerchief when coughing or sneezing (true) |
| KP05 | When this virus spreads in an area, people should stay home as much as possible to reduce the spread of this disease (true) |
| KP06 | Infected person should wear mask to reduce the spread of this virus (true) |
|  | **Section-3: Questions related to attitude towards COVID-19 (only for online survey)** |
| A1 | Do you think that this new coronavirus will finally be under control completely? |
| A2 | “This virus is created by humans” - this kind of discourse is heard. Do you believe that it is true? |

The questionnaire had three main sections: Section-1 asked about the source of information regarding COVID-19 or SARS-CoV-2. Section-2 (knowledge section) had two subdivisions: Section-2A (questions related to the knowledge of disease) and Section-2B (questions related to the knowledge of preventive practices). Section-3 (attitude section) was used only for the online survey. In Section-2A, there were 14 questions (KD01 to KD14) related to the Knowledge of Disease that included signs/symptoms, mode of spread, outcome of the disease etc. In Section-2B, there were six questions (KP01 to KP06) related to the Knowledge of Preventive practices. In Section-3, there were two attitude questions (A1 and A2). Options to the knowledge questions were a) true, b) false, and c) don’t know. Options to the attitude questions were a) yes, b) no, and c) don’t know. A correct response to a knowledge question was assigned 1 point and an incorrect/don’t know response was assigned 0 point. Total score of Knowledge of Disease (KD) and Knowledge of Preventive practices (KP) ranged from 0 to14 and 0 to 6, respectively. Therefore, the total knowledge score ranged from 0-20. A higher score indicated a higher knowledge level. The Cronbach’s alpha coefficient of the knowledge questionnaire was 0.74 in the current study, indicating acceptable internal consistency [10, 24].

### 2.4. Statistical Analysis

Data analysis was primarily done with IBM SPSS Statistics 20. We divided the survey participants into three main groups as “phone”, “online non-medical”, and “online medical”. Frequencies and percentages were used to describe the responses to the questions and sociodemographic characteristics of the participants. Chi-square test for independence and Mann-Whitney U test were used for comparison between different groups wherever appropriate. Correlations among KD, KP, and total knowledge scores were investigated with Pearson product-moment correlation. Binary or multinomial logistic regression was used as appropriate to identify factors associated with total knowledge score and attitudes. The statistical significance level was set at *P* < 0.05 (two-sided). Benjamini-Hochberg correction of *P* values was done as appropriate in case of multiple testing and statistical significance was determined using a false discovery rate (FDR) of 0.05.

The R package “anesrake” version 0.80 was used for raking (also known as iterative post-stratification) the phone survey data for different purposes [25]. The unadjusted phone survey data was weighted (adjusted) to match the socio-demographic distributions (age, gender, education, lived mostly in last four months, financial condition of the family) of the online non-medical survey data for comparison of the phone and online survey methods. For population-level estimation, the unadjusted phone survey data was adjusted to match the socio-demographic distributions of Bangladeshi adult population (as of 2011 Bangladesh national census data, given in the supplementary methods).

## 3. Results

There were 1427 phone participants. All of them heard about COVID-19 except one. In the online survey, we had a total of 1521 participants. 98.6% (1500) of them heard about the disease. From these 1500 online participants, 19 were excluded because of being less than 18 years old. Two more participants were excluded because they reported their education level as not more than grade 12 (higher secondary school – HSC) but reported themselves as doctor/nurse/medical student. The results presented hereafter are based on the final dataset of 1426 phone participants and 1479 online participants. Among these 1479 online participants, 382 were medical and the remaining 1097 were non-medical.

The administrative division-wise distribution of the participants is given in Table S1 and Fig. S1. We had participants from all the divisions of Bangladesh in both phone and online surveys although most of them were from the two major divisions Chattogram and Dhaka. The other socio-demographic characteristics are shown in Table 2. Chi-square test for independence showed that all the characteristics differed significantly between participant groups. The participants of the phone survey were less educated and from poorer families compared to the online participants. A significantly higher portion of the online participants had been living in urban/town areas (vs. rural) for the last four months compared to the phone survey participants. Also, the online survey had comparatively more young participants than phone survey. Both phone and online survey participants mentioned television and internet as major sources of information of COVID-19 (Fig. 1a).

**Table 2.**
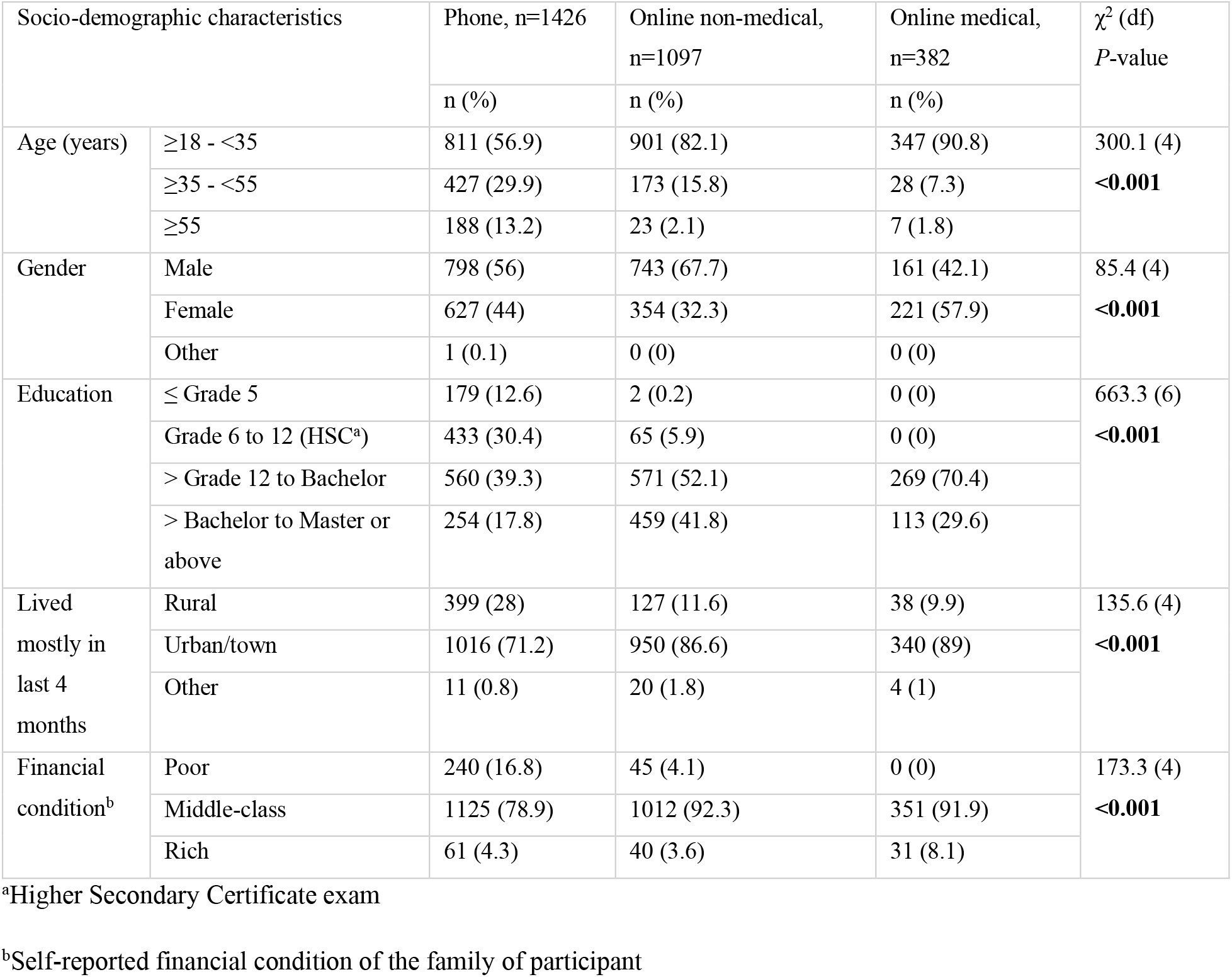
Socio-demographic characteristics of the participants

| Socio-demographic characteristics | | Phone, n=1426 | Online non-medical,<br>n=1097 | Online medical,<br>n=382 | $\chi^2$ (df)<br>P-value |
| --- | --- | --- | --- | --- | --- |
|  |  | n (%) | n (%) | n (%) |  |
| Age (years) | ≥18 - <35 | 811 (56.9) | 901 (82.1) | 347 (90.8) | 300.1 (4) |
|  | ≥35 - <55 | 427 (29.9) | 173 (15.8) | 28 (7.3) | **<0.001** |
|  | ≥55 | 188 (13.2) | 23 (2.1) | 7 (1.8) |  |
| Gender | Male | 798 (56) | 743 (67.7) | 161 (42.1) | 85.4 (4) |
|  | Female | 627 (44) | 354 (32.3) | 221 (57.9) | **<0.001** |
|  | Other | 1 (0.1) | 0 (0) | 0 (0) |  |
| Education | ≤ Grade 5 | 179 (12.6) | 2 (0.2) | 0 (0) | 663.3 (6) |
|  | Grade 6 to 12 (HSC<sup>a</sup>) | 433 (30.4) | 65 (5.9) | 0 (0) | **<0.001** |
|  | > Grade 12 to Bachelor | 560 (39.3) | 571 (52.1) | 269 (70.4) |  |
|  | > Bachelor to Master or above | 254 (17.8) | 459 (41.8) | 113 (29.6) |  |
| Lived mostly in last 4 months | Rural | 399 (28) | 127 (11.6) | 38 (9.9) | 135.6 (4) |
|  | Urban/town | 1016 (71.2) | 950 (86.6) | 340 (89) | **<0.001** |
|  | Other | 11 (0.8) | 20 (1.8) | 4 (1) |  |
| Financial condition<sup>b</sup> | Poor | 240 (16.8) | 45 (4.1) | 0 (0) | 173.3 (4) |
|  | Middle-class | 1125 (78.9) | 1012 (92.3) | 351 (91.9) | **<0.001** |
|  | Rich | 61 (4.3) | 40 (3.6) | 31 (8.1) |  |

<sup>a</sup>Higher Secondary Certificate exam<sup>b</sup>Self-reported financial condition of the family of participant

**Fig. 1.**
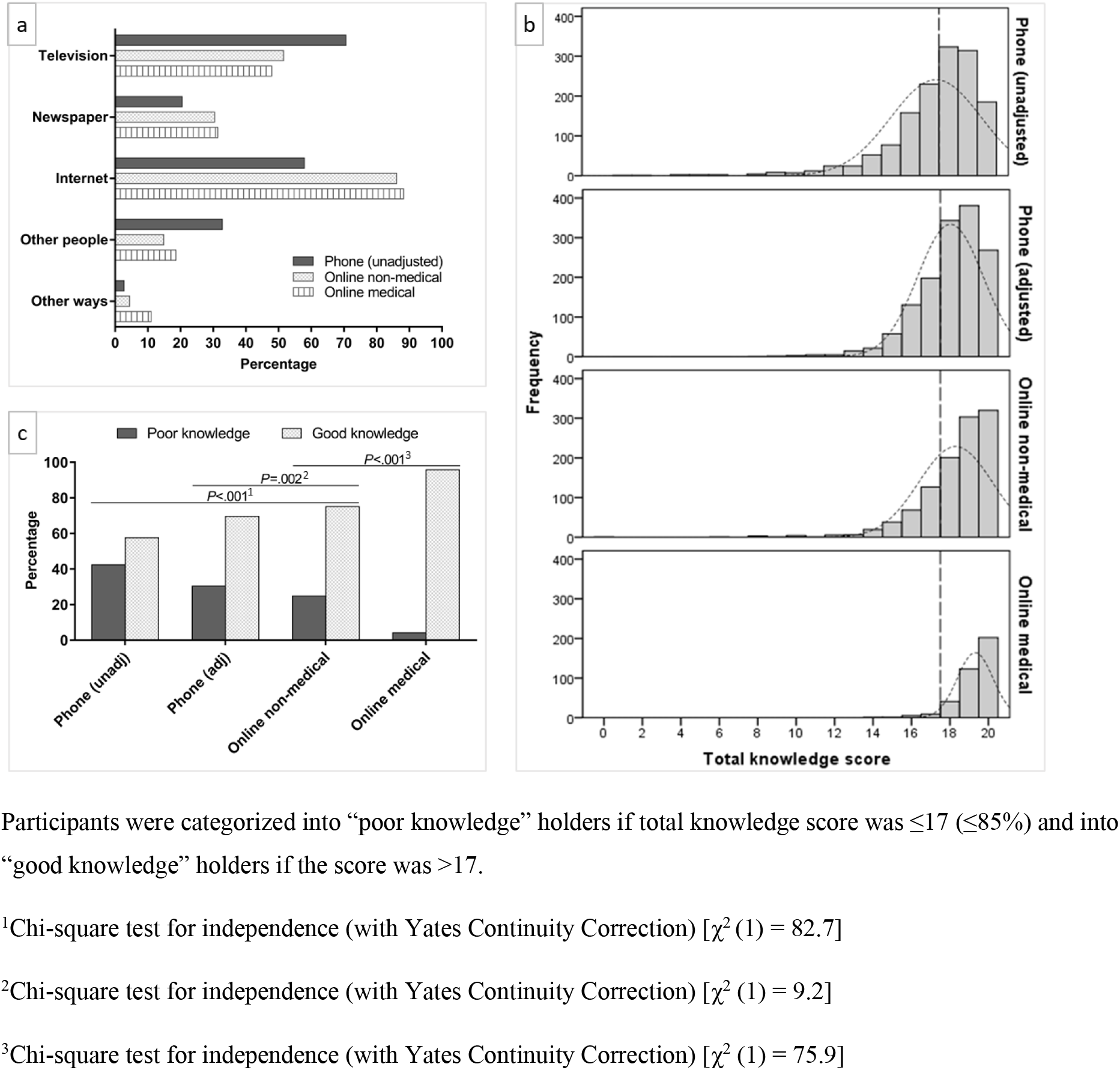
Sources of information according to the participant groups (1a), distribution of total knowledge scores (1b), and percentage of poor vs. good knowledge holders in each participant group (1c)

The correct response rates to the 20 knowledge questions ranged 54.3-99.1%, 64-99.6%, 71.1-99.9%, and 88-100% in the phone (unadjusted), phone (adjusted to online non-medical), online non-medical, and online medical participants, respectively (Table 3). Chi-square test for independence showed that the correct response rates differed significantly for 11 out of 14 KD questions between the unadjusted phone survey data and online non-medical survey data. In general, for questions having significantly different correct response rates, the rates were higher in online survey compared to that of the phone survey, except for two questions (KD02 and KD10). However, the number of KD questions having significantly different correct response rates dropped to six when the phone survey data was adjusted to the socio-demographic characteristics of the online non-medical participant group. In addition, for 11 out of 14 KD questions, correct response rates were significantly higher for the online medical group than online non-medical. This suggests that the knowledge of disease was the highest in the online medical group which was followed by the online non-medical group. The phone survey group was the least knowledgeable about the disease even after adjusting to the online non-medical group. Despite this difference in KD, correct response rates in the KP questions showed less difference except for one (KP01) question.

**Table 3.** Percentage of the correct response to each question of the knowledge questionnaire

|  | 1 | 2 | 3 | 4 | 1 vs. 3 |  | 2 vs. 3 |  | 3 vs. 4 |  |
| --- | --- | --- | --- | --- | --- | --- | --- | --- | --- | --- |
| Items | Phone<br>(unadjusted)<br>n=1426 | Phone<br>(adjusted to<br>online non-medical)<sup>a</sup><br>n=1426 | Online non-medical<br>n=1097 | Online<br>medical<br>n=382 | Chi-square test<sup>b</sup> |  | Chi-square test<sup>b</sup> |  | Chi-square test<sup>b</sup> |  |
| | % | % | % | % | $\chi^2$ | *P*<sup>c</sup> | $\chi^2$ | *P*<sup>c</sup> | $\chi^2$ | *P*<sup>c</sup> |
| Section-1A: Questions related to the knowledge of the disease (KD) |  |  |  |  |  |  |  |  |  |  |
| KD01 | 54.3 | 67.0 | 71.1 | 94.2 | 73.5 | **<0.001** | 4.7 | 0.055 | 84.6 | **<0.001** |
| KD02 | 89.7 | 89.1 | 86.0 | 93.2 | 7.9 | **0.011** | 5.2 | **0.043** | 13.2 | **<0.001** |
| KD03 | 87.8 | 87.3 | 86.2 | 94.5 | 1.2 | 0.416 | 0.5 | 0.620 | 18.0 | **<0.001** |
| KD04 | 93.4 | 96.6 | 96.6 | 99.2 | 12.4 | **0.001** | 0.0 | >0.99 | 6.3 | **0.026** |
| KD05 | 86.5 | 89.5 | 92.6 | 96.9 | 23.1 | **<0.001** | 7.0 | **0.018** | 7.9 | **0.012** |
| KD06 | 56.6 | 64.0 | 77.8 | 88 | 122.5 | **0.001** | 55.0 | **<0.001** | 18.1 | **<0.001** |
| KD07 | 64.1 | 75.3 | 77.2 | 94.2 | 50 | **<0.001** | 1.1 | 0.423 | 53.6 | **<0.001** |
| KD08 | 86.8 | 89.8 | 92.3 | 95.5 | 19.1 | **<0.001** | 4.4 | 0.064 | 4.1 | 0.076 |
| KD09 | 84.4 | 91.4 | 95.3 | 99.5 | 74.7 | **<0.001** | 13.9 | **0.001** | 13.1 | **<0.001** |
| KD10 | 82.3 | 80.9 | 76.2 | 88 | 14 | **0.001** | 8.0 | **0.012** | 23.1 | **<0.001** |
| KD11 | 96.6 | 98.2 | 97.9 | 99 | 3.5 | 0.102 | 0.1 | 0.929 | 1.2 | 0.408 |
| KD12 | 96.6 | 98.9 | 99.1 | 99.2 | 15.6 | **<0.001** | 0.0 | >0.99 | 0.0 | >0.99 |
| KD13 | 83.6 | 93.9 | 97.6 | 100 | 130.7 | **<0.001** | 19.3 | **<0.001** | 7.9 | **0.011** |
| KD14 | 93.1 | 95.7 | 94.3 | 98.7 | 1.4 | 0.386 | 2.2 | 0.225 | 11.4 | **0.002** |
| Section-1B: Questions related to the knowledge of the preventive practices (KP) |  |  |  |  |  |  |  |  |  |  |
| KP01 | 86.9 | 90.7 | 95.0 | 96.9 | 45 | **<0.001** | 16.1 | **<0.001** | 1.9 | 0.276 |
| KP02 | 98.9 | 98.7 | 99.1 | 99 | 0.1 | 0.936 | 0.4 | 0.680 | 0.0 | >0.99 |
| KP03 | 98.4 | 98.9 | 98.5 | 98.4 | 0 | >0.99 | 0.6 | 0.610 | 0.0 | >0.99 |
| KP04 | 99.1 | 99.6 | 99.9 | 99.7 | 6.1 | **0.026** | 0.8 | 0.504 | 0.0 | >0.99 |
| KP05 | 99.0 | 99.2 | 99.2 | 99.5 | 0.4 | >0.99 | 0.0 | >0.99 | 0.2 | >0.99 |
| KP06 | 96.8 | 97.7 | 97.5 | 97.6 | 0.8 | 0.512 | 0.0 | >0.99 | 0.0 | >0.99 |

<sup>a</sup>The participants of the unadjusted phone survey dataset were weighted by raking to match the sociodemographic distributions of the online non-medical survey dataset
<sup>b</sup>Chi-square test for independence (with Yates Continuity Correction) (df=1)
<sup>c</sup>Benjamini-Hochberg Adjusted *P* values. *P* values that are significant using an FDR of 0.05 are made bold.

Participants were categorized into “poor knowledge” or “good knowledge” holders based on the total knowledge score achieved (≤17 and >17-20 respectively) (Fig. 1b and 1c). The percentages of good knowledge holders in the phone (unadjusted), phone (adjusted to online non-medical), online non-medical, and online medical groups were 57.6%, 69.6% 75.1%, and 95.8%, respectively (Fig. 1c). The total knowledge scores based on sociodemographic characteristics and sources of COVID-19 information are shown in Table 4. Online medical participants achieved the highest total knowledge score (median 20, IQR 19-20) (Table 4). It was followed by the online non-medical group (median 19, IQR 18-20). The phone participants scored the least (median 18 [IQR 16-19] and median 18 [IQR 17-19] for unadjusted and adjusted data, respectively). The median scores for almost all categories of sociodemographic characteristics and sources of information were higher in online non-medical group compared to that of the phone participant group with most of the differences being statistically significant.

**Table 4.**
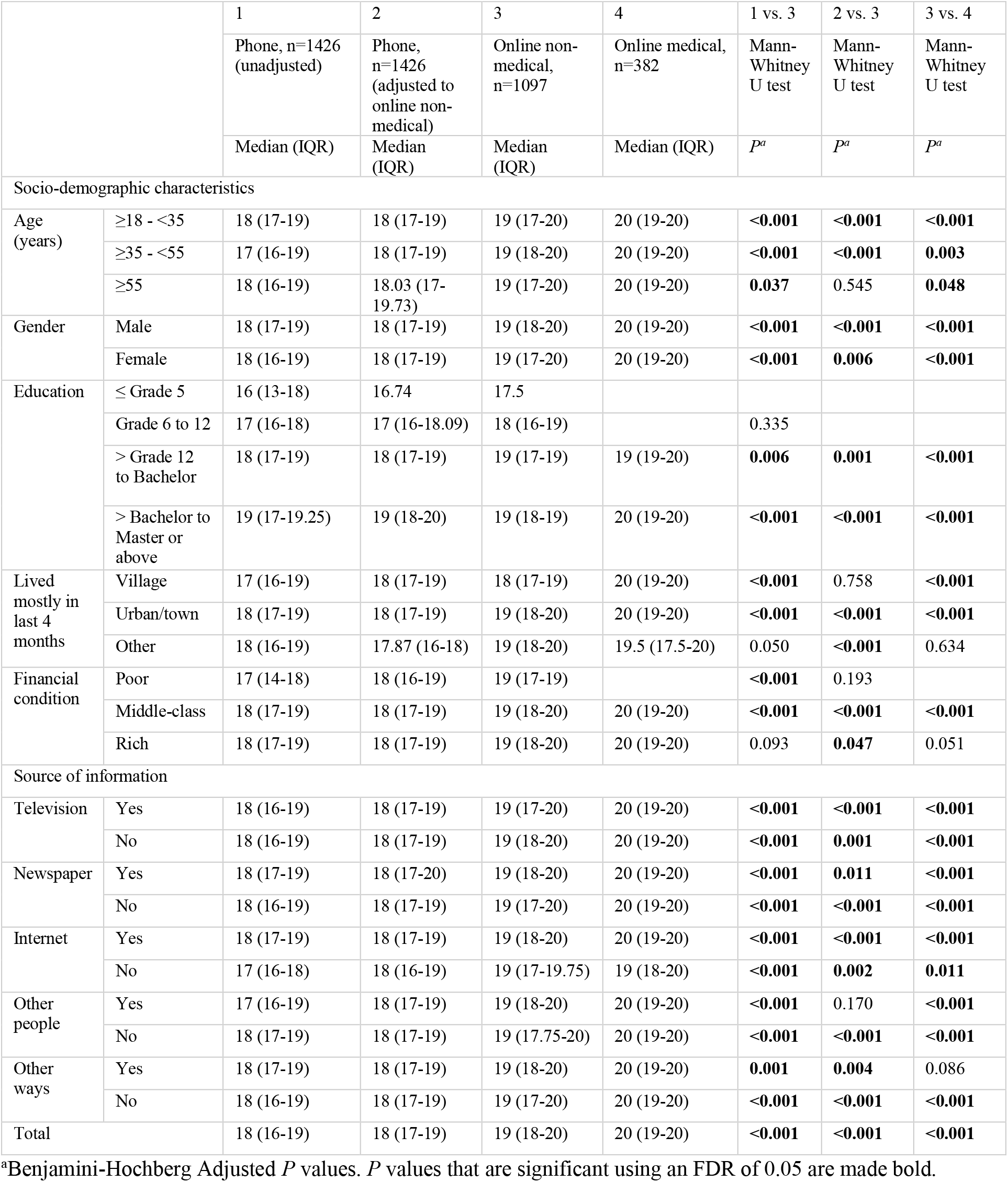
Total knowledge score according to the sociodemographic characteristics and source of information

Pearson product-moment correlation coefficient showed that there was a positive association of medium strength (coefficient 0.33) between KD score and KP score (Table 5). Although both KD and KP had strong correlation with total knowledge score, the KD had a nearly perfect correlation with the total knowledge score with a coefficient of 0.98 and thereby explaining 96.2% of the variance in the total knowledge score. Taken together with the high correct response rates to five of the six KP questions (as shown in Table 3), it suggests that most participants had good knowledge of the preventive practices irrespective of their knowledge level of the disease.

**Table 5.** Pearson product-moment correlations between different scores

| Scale | KD score | KP score | Total knowledge score |
| --- | --- | --- | --- |
| KD score | - | 0.33* | 0.98* |
| KP score |  | - | 0.51* |
| Total knowledge score |  |  | - |

\* $P < 0.001$

Binary logistic regression analysis of the combined dataset of phone (unadjusted) and online non-medical participants showed that, male gender (vs. female), higher education (vs. ≤ grade 12), living in town for the last four months (vs. living in rural areas), middle-class/rich financial condition (vs. poor), and internet as a source of information (vs. not as a source) were positively associated with having good knowledge (vs. poor knowledge) (Table 6).

**Table 6.** Results of binary logistic regression on factors predicting the likelihood of having good knowledge

| Factors |  | B | S.E. | Wald | df | P | Odds Ratio | 95% C.I. for Odds Ratio |  |
| --- | --- | --- | --- | --- | --- | --- | --- | --- | --- |
|  |  |  |  |  |  |  |  | Lower | Upper |
| Age | ≥35 to <55 vs ≥18 to <35 | 0.09 | 0.12 | 0.58 | 1 | 0.445 | 1.10 | 0.87 | 1.39 |
|  | ≥55 vs ≥18 to <35 | 0.25 | 0.17 | 1.99 | 1 | 0.158 | 1.28 | 0.91 | 1.80 |
| Gender | Male vs. Female | 0.31 | 0.09 | 11.12 | 1 | **0.001** | 1.36 | 1.14 | 1.63 |
| Education | >HSC to Bachelor vs. ≤HSC | 0.58 | 0.13 | 19.98 | 1 | **<0.001** | 1.79 | 1.39 | 2.31 |
|  | >Bachelor vs. ≤HSC | 1.22 | 0.15 | 69.80 | 1 | **<0.001** | 3.38 | 2.54 | 4.49 |
| Lived in last 4 months | Town vs. Rural | 0.26 | 0.11 | 5.40 | 1 | **0.020** | 1.30 | 1.04 | 1.62 |
| Financial condition | Middle-class/Rich vs. Poor | 0.34 | 0.15 | 5.13 | 1 | **0.024** | 1.41 | 1.05 | 1.90 |
| Source of information | Television (yes vs. no) | 0.04 | 0.10 | 0.12 | 1 | 0.727 | 1.04 | 0.85 | 1.27 |
|  | Newspaper (yes vs. no) | 0.16 | 0.12 | 1.92 | 1 | 0.166 | 1.18 | 0.93 | 1.49 |
|  | Internet (yes vs. no) | 0.58 | 0.11 | 25.78 | 1 | **<0.001** | 1.79 | 1.43 | 2.24 |
|  | Other people (yes vs. no) | -0.14 | 0.11 | 1.65 | 1 | 0.200 | 0.87 | 0.70 | 1.08 |
|  | Other ways (yes vs. no) | 0 | 0.27 | 0 | 1 | >0.99 | 1.00 | 0.60 | 1.69 |
| Constant |  | -1.09 | 0.19 | 31.68 | 1 | **<0.001** | 0.34 |  |  |

*P* values less than 0.05 are made bold

Responses to the attitude questions are shown in Table 7 (responses by sociodemographic characteristics and source of information are shown in Table S2 and Table S3). Among the online non-medical and online medical participants, 61.1% and 56% respectively were optimistic that this new coronavirus will finally be controlled completely. Multinomial logistic regression analysis showed that for every unit increase in the total knowledge score, the odds of the participants responding “no” (compared to responding “yes”) increased by a factor of 1.19 (95% CI 1.04 to 1.36), all other factors being equal (Table 8). Similarly, the odds of a participant responding “no” (compared to “yes”) was 2.95 times higher (95% CI 1.28 to 6.78) for someone who reported the internet as a source of information than for a participant who did not, all other factors being equal. Interestingly, females had higher odds of responding “don’t know” (compared to “yes” or “no”) to this attitude question than males.

**Table 7.** Responses to the attitude questions

| Attitude questions |  | Online non-medical, n=1097 |  |  | Online medical, n=382 |  |  |
| --- | --- | --- | --- | --- | --- | --- | --- |
|  |  | Yes | No | Don't know | Yes | No | Don't know |
| A1. Do you think that this new coronavirus will finally be under control completely? | n (%) | 670 (61.1) | 119 (10.8) | 308 (28.1) | 214 (56) | 63 (16.5) | 105 (27.5) |
| A2. "This virus is created by humans" - this kind of discourse is heard. Do you believe that it is true? | n (%) | 210 (19.1) | 280 (25.5) | 607 (55.3) | 69 (18.1) | 138 (36.1) | 175 (45.8) |

**Table 8.**
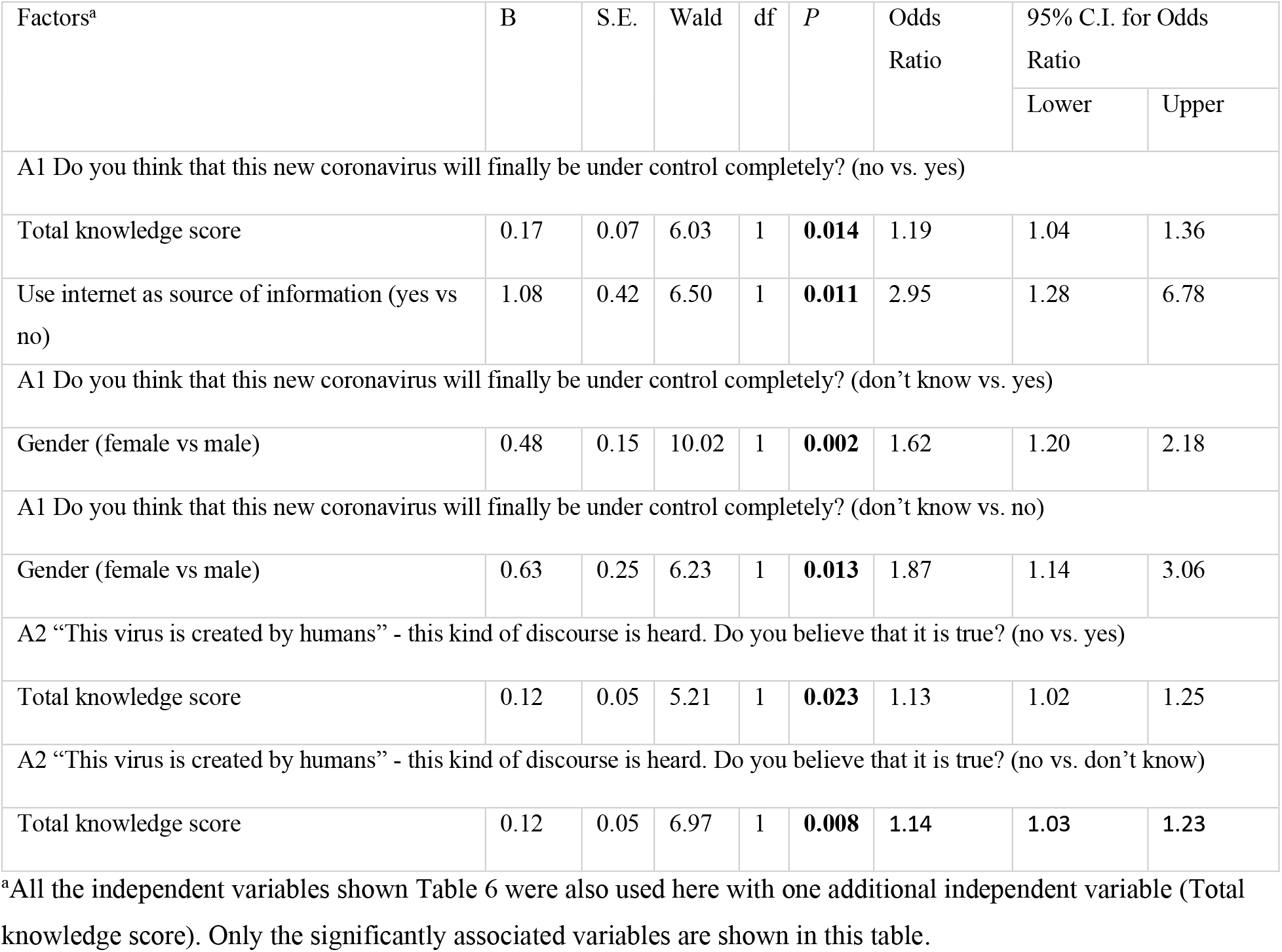
Multinomial logistic regression analysis on factors predicting the likelihood of different responses of attitude questions

In case of attitude question A2 (“This virus is created by humans” - this kind of discourse is heard. Do you believe that it is true?), only 25.5% and 36.1% of the online non-medical and online medical participants, respectively, believed that the virus was not created by humans (Table 7). Most of the participants responded “don’t know” to this attitude question. Multinomial logistic regression analysis showed that for every unit increase in the total knowledge score, the odds of the participants responding “no” increased by a factor of 1.13 or 1.14 (compared to responding “yes” or “don’t know”, respectively), all other factors being equal (Table 8).

Finally, we adjusted the original phone survey data to the sociodemographic distributions (gender, education, rural vs. urban/town, and poor vs. middle-class/rich) of the Bangladeshi adult population in an effort to estimate the correct response rates to the knowledge questions at population level (Table 9). It estimated that less than 50% of the adult population know that some COVID-19 patients might have no symptoms, diarrhea is a symptom of COVID-19, and this disease cannot be completely prevented by any medications. The estimated average total knowledge score was 15.85 (SD 3.14) with a median score of 17 (IQR 14-18). Only 32.6% (Clopper-Pearson 95% CI 30.1-35.2%) of the population was estimated to have good knowledge.

**Table 9.** Population-level estimates of the correct response rates to the knowledge questions

| Questions related to the knowledge of disease |  |  |  | Questions related to the knowledge of preventive practices |  |  |  |
| --- | --- | --- | --- | --- | --- | --- | --- |
| Items | Estimated correct response rate (%) | Clopper-Pearson 95% confidence interval |  | Items | Estimated correct response rate (%) | Clopper-Pearson 95% confidence interval |  |
|  |  | Lower (%) | Upper (%) |  |  | Lower (%) | Upper (%) |
| KD01 | 31.2 | 28.6 | 33.8 | KP01 | 77.5 | 75.2 | 79.8 |
| KD02 | 91.3 | 89.6 | 92.8 | KP02 | 98 | 97.1 | 98.7 |
| KD03 | 88.2 | 86.3 | 89.9 | KP03 | 96 | 94.8 | 97 |
| KD04 | 86.8 | 84.9 | 88.6 | KP04 | 96.4 | 95.2 | 97.3 |
| KD05 | 79.2 | 76.8 | 81.3 | KP05 | 97.2 | 96.1 | 98 |
| KD06 | 37.5 | 34.8 | 40.2 | KP06 | 94.9 | 93.6 | 96.1 |
| KD07 | 41.4 | 38.7 | 44.1 |  |  |  |  |
| KD08 | 78.6 | 74.5 | 79.1 |  |  |  |  |
| KD09 | 66.9 | 64.3 | 69.5 |  |  |  |  |
| KD10 | 82 | 79.8 | 84.1 |  |  |  |  |
| KD11 | 90.7 | 89 | 92.2 |  |  |  |  |
| KD12 | 90.9 | 89.2 | 92.4 |  |  |  |  |
| KD13 | 61.4 | 58.7 | 64 |  |  |  |  |
| KD14 | 85.2 | 83.1 | 87 |  |  |  |  |

## 4. Discussion

Bangladesh is a lower-middle-income country and one of the most densely populated countries in the world. Apparently, like many other countries, it is struggling to combat COVID-19. Nearly all countries are relying heavily on non-pharmacological interventions against COVID-19. Success in this battle is partly determined by the knowledge and behavioral changes of the general population as well as how effectively these non-pharmacological control measures are being implemented by the authorities and abided by the citizens [26]. Therefore, it is imperative to know about the knowledge and attitudes of the population towards COVID-19 to develop effective strategies. Given the social distancing measures in place in most countries, the studies quantifying KAP of the population were mostly conducted through internet with some exceptions [26]. Survey through the internet may quantify the true KAP of the population of countries where a high proportion of the population use internet (e.g., 95% and 87% of the population of the United Kingdom and the United States use internet, respectively [World Bank – 2017]) [23]. However, online survey may not be able to quantify the true KAP of the population of countries where access to the internet is limited and may result in overestimation of the KAP. In addition, because of inadequate samples from underrepresented population groups (e.g., poor, less uneducated, and rural people), it becomes difficult to adjust the online survey data to the general population of the respective countries with poor internet access [21, 22]. In the present study, we primarily addressed two key issues. First, we tried to determine whether there is a significant difference in the estimated COVID-19 knowledge level determined by the online and phone survey methods given the same questionnaire. Second, we tried to quantify the knowledge of COVID-19 in Bangladeshi adult population.

In our study, the sociodemographic characteristics were significantly different between the phone and online participants. The phone survey method was able to include more participants from poor, rural, and less educational backgrounds compared to the online survey. In fact, there were only a few participants in the online survey in some socio-demographic groups (e.g., education up to grade five) making it difficult to adjust the online dataset to the population level. To compare the estimated knowledge level by the phone survey and that by the online survey, we adjusted the phone survey data to the sociodemographic characteristics of the online survey dataset. We found that, the total knowledge score was still significantly higher in the online non-medical participants than that of phone survey participants in most of the socio-demographic groups. Although investigating the factors responsible for the higher estimated knowledge level in the online survey was out of the scope of our current study, online participants probably had more opportunity to know the correct answers from others or by searching elsewhere than the phone survey participants who had to respond immediately to the survey questions during the phone calls. It is noticeable from our study that the total knowledge score did not differ significantly between the adjusted phone survey dataset and online non-medical dataset for the elderly (≥55 years), poor, and rural people. It suggests that, although these people are less likely to be accessed by the online surveys in countries like Bangladesh, the knowledge estimates will be similar by both phone and online survey methods for these subgroups of the population. However, as these people are likely to be underrepresented in online surveys and more educated, urban, younger, and financially solvent people are likely to be overrepresented, we therefore advise caution in generalizing the unadjusted online survey results to the general population particularly in countries with limited internet access for the population.

In the present study, inclusion of participants from medical backgrounds in the online survey allowed us to choose an informed cut-off score for dividing the participants into poor or good knowledge holders rather than choosing an arbitrary score as in some other COVID-19 KAP studies [21, 10]. We assumed that most of the medical participants should have good knowledge of COVID-19. Then we examined the distribution of total knowledge scores (Fig. 1b) and decided to use a score of >17 (out of 20) as a cut-off score to categorize participants as good knowledge holders as it covered 95.8% of the medical participants as good knowledge holders. Based on this criterion, we found that 75.1% of the online non-medical participants had good knowledge which was significantly higher than that of phone survey participants.

In Fig. 2, we tried to summarize the timeline of all the major COVID-19 related events that occurred in Bangladesh (adapted and updated from [27]). We also highlighted the timeline of two other online surveys conducted previously in Bangladesh so that we can meaningfully compare our online survey results with those studies [21, 22]. The first study by Haque, T et al. [21] was done at the very beginning of the COVID-19 spread in Bangladesh. The study ended on March 30, when Bangladesh had total confirmed cases of 49 (deaths 5). The second study by Rahman, A et al. [22], was done for four days that ended on April 10, at the time when the total number of confirmed cases started to rise rapidly (from 164 to 426 in four days). The online survey of the present study was done approximately 1 week later from that study when the total number of confirmed cases was 2144 (April 18). Although the questionnaires used by those two previous studies and in our study are different, all these studies should yield comparable results as the questionnaires were essentially developed based on the information provided by the leading authorities (WHO, IEDCR) for the general public. The average total knowledge scores were 76%, 90.7%, and 91.5%, respectively. The percentages of good knowledge holders were 54.9%, 86.2%, and 75.1%, respectively (although this is prone to more variation than the average knowledge score as the defining scores for “good knowledge” were somewhat arbitrary). Therefore, it is reasonable to claim that there has been a clear improvement in the knowledge level among the online participants. However, to what extent this improvement applies to the huge portion of the Bangladeshi population who do not have access to the internet, remains unclear as we could not find any previous data to compare our phone survey data. Unfortunately, the improvement might not be as dramatic as online participants. There could have been a lot of factors associated with the improvement of knowledge level of the online participants. Fig. 2 shows the trends of “corona”-related news on two popular Bangladeshi online news portals and the Google Trends for search term “coronavirus” from Bangladesh. The news trends rapidly raised soon after the initial declaration of the COVID-19 cases on March 8 indicating that people were getting increasing amount of coronavirus-related news from the news portals as well as sharing of the news on social media. Some major unprecedented steps taken by the authorities also followed soon. All these factors could gradually have made people more interested to actively learn about the disease as evident in the Google Trends. This is also supported by the findings of a positive association of good COVID-19 knowledge level with higher education and use of internet as a source of information in our study. However, it is extremely difficult to quantify the relative contribution of all these factors to the improvement of the knowledge level. An online survey conducted in China approximately one month after the initial case detection in Wuhan, found a similar knowledge level among the participants (average total knowledge score of 90%) [10].

**Fig. 2.**
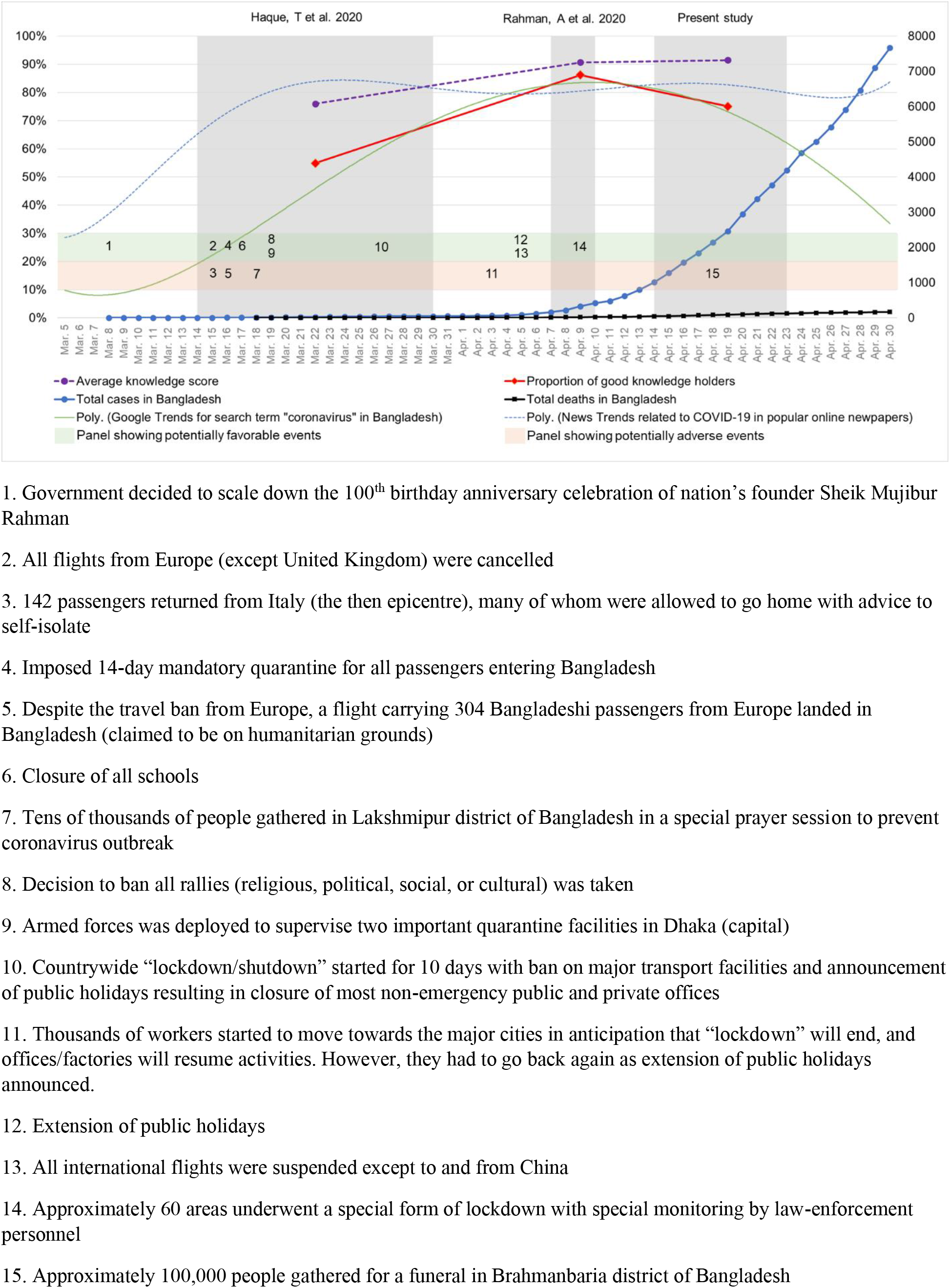
Timeline of all major events related to COVID-19 in Bangladesh

In the study from China mentioned above, 90.8% of the online respondents were optimistic that the COVID-19 will finally be successfully controlled. Furthermore, higher COVID-19 knowledge score was positively associated with higher likelihood of being optimistic about this [10]. Apparently, compared to many other countries, China has been successful in controlling the local epidemic approximately within two months of the outbreak despite being the initial epicenter of COVID-19. Contrasting to this, and possibly a warning finding in our study, only 61.1% of the online non-medical participants had optimistic attitudes towards final control of COVID-19. The medical participants were even less optimistic (56%). Rahman. A et al. reported even lower proportion of online respondents having optimistic attitudes (41.7%) [22]. Furthermore, in our combined dataset of phone (unadjusted) and online non-medical participants, we found that higher total knowledge score was positively associated with having negative attitude towards the final control of COVID-19. If we look back to the timeline presented in Fig. 2 and see the potentially major adverse events that happened in Bangladesh (see the paper by [27] for detailed discussion) that were in stark contrast to the known containment strategies, it is tempting to associate these events to the negative feelings towards the final success. One might argue that Bangladesh could suffer devastating consequences if more effective and coordinated approaches are not taken and people continue to defy social distancing advice. However, as pointed out by [27], it is extremely challenging in Bangladesh.

Our study estimated that the average total knowledge score would be approximately 79.3% (15.85/20) in the general Bangladeshi adult population with an estimated 32.6% of the population having good knowledge. This finding is alarming for the country and appropriate educational public health measures must be taken to improve the knowledge level so that the people can have better chance of protecting themselves.

One limitation of our study was the sampling techniques. As we had to rely on the authors’ network for recruiting participants for phone survey, it suffered selection bias and we could not obtain a nationally representative sample for adult population. However, as we obtained good number of participants from each sociodemographic category and adjusted the dataset to national census data, this issue should be less problematic, and our population-level estimates should be fairly representative. The second limitation worth mentioning is that we merely assessed the knowledge level of preventive practices. Thus, the knowledge level found for the preventive practice section does not necessarily reflect the actual physical behavior adopting those preventive practices. It is reasonable to argue that the proportion of the population adopting those preventive practices could be less. For example, our population level estimates showed that 98% of the population know the information that “washing hands frequently and thoroughly with soap or alcohol-based hand sanitizers reduces the risk of being infected by the virus”. But in practice, proportion of the population washing hands more frequently could be less. A third limitation was that we could not quantify the financial condition of the participants with any formal scale to limit the phone call duration. Therefore, we had to rely on the “self-reported” financial condition.

## 5. Conclusion

Our findings suggest that online survey might overestimate knowledge level compared to phone survey, although there appears to be no difference for elderly, poor, and rural people. We also showed evidence supporting a rapidly increasing COVID-19 knowledge level among the online participants. Male gender, higher education, living in town/urban areas, good financial condition, and use of internet were positively associated with higher knowledge. However, higher knowledge was associated with having less confidence in the final control of COVID-19. Our population-level estimates showed that only 32.6% had good knowledge.

## Data Availability

Survey data are available from the corresponding author on request.

## Ethical approval

The study was approved by the Ethical Review Committee of Chittagong Medical College, Chattogram, Bangladesh.

## Acknowledgement

The authors would like to express great appreciation to Mahadi Shaded (City University of Hong Kong), Md Arzan Hosen (First Capital University of Bangladesh), and Umme Hafsa (International Islamic University Chittagong, Bangladesh) for their comments on the study design and contribution to data collection.

## Data sharing

Survey data are available from the corresponding author on request.

## Supplementary material to

### Supplementary methods

<u>Raking the phone survey dataset to adjust to the online nonmedical dataset</u>

- Complete convergence was achieved after 22 iterations in anesrake version 0.80

<u>Raking the phone survey dataset to adjust to the adult population level</u>

- Target population level proportions used for raking:
  - Gender
    - Male: 0.527
    - Female: 0.473
  - Education
    - ≤ Grade 5: 0.734
    - Grade 6 to grade 12 (HSC): 0.230
    - ≥ HSC to Bachelor: 0.024
    - ≥ Bachelor to Master or above: 0.012
  - Living area
    - Rural: 0.634
    - Urban/town: 0.366
  - Economic condition
    - Poor: 0.218
    - Middle-class/rich: 0.782
- Complete convergence was achieved after 608 iterations

<u>Google Trends for search term “coronavirus”</u>

- Data on Google Trends for the search term “coronavirus” was extracted from https://trends.google.com/trends/explore?date=2020-03-05%202020-04-30&geo=BD&q=coronavirus
- The Polynomial trendline (Order 6) that is shown in Fig. 2 of the manuscript was created with MS Excel

<u>News Trends related to COVID-19 in popular online newspapers</u>

Two popular online news portals (www.prothomalo.com, www.jagonews24.com) were used for this purpose. We first calculated the total number of reports in both news portals with keyword “corona” on each day from March 5 to April 30, 2020. Then the highest total was considered as 100% and the other total number of reports for each day was then expressed as percentage relative to the highest number. The Polynomial trendline (Order 6) that is shown in Fig. 2 of the manuscript was created with MS Excel

### Supplementary results

**Table S1.**
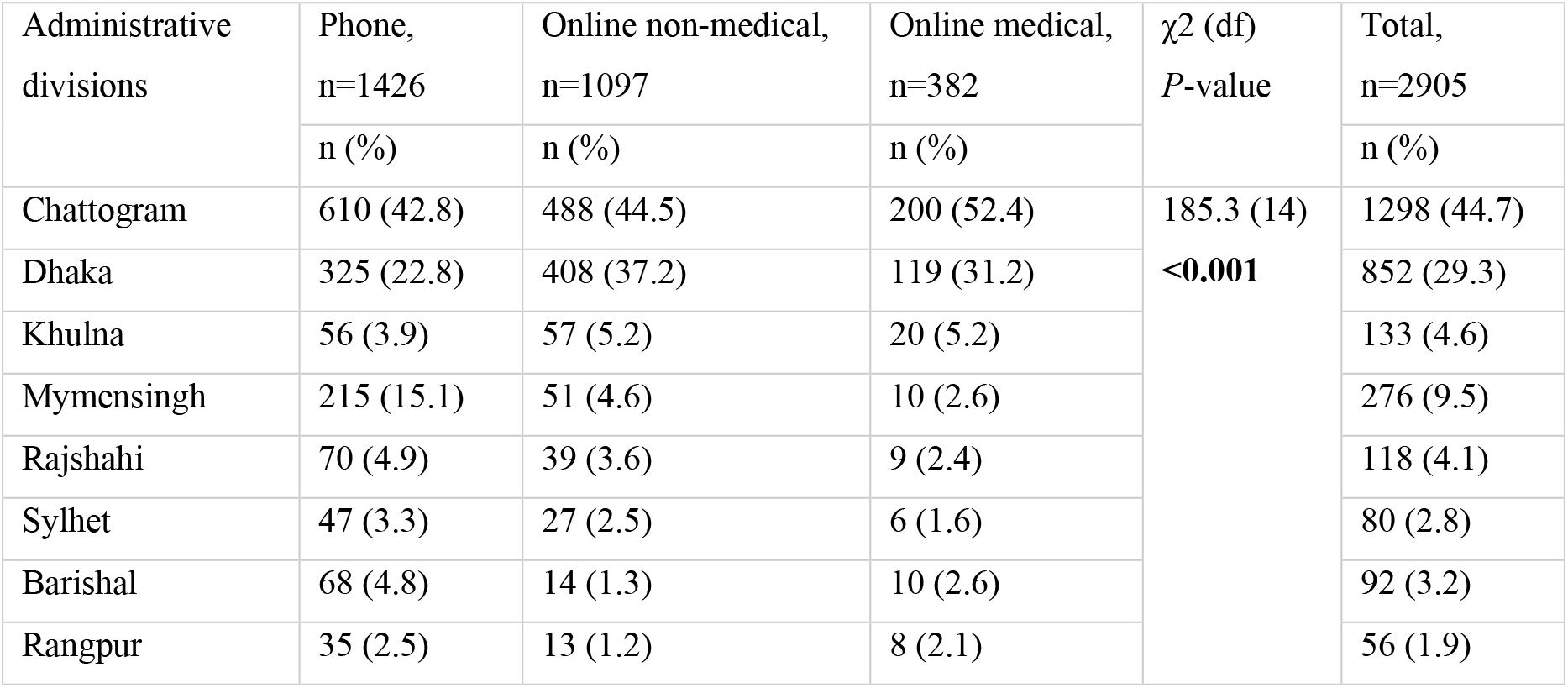
Distribution of the participants according to the eight administrative divisions of Bangladesh

**Fig. S1.**
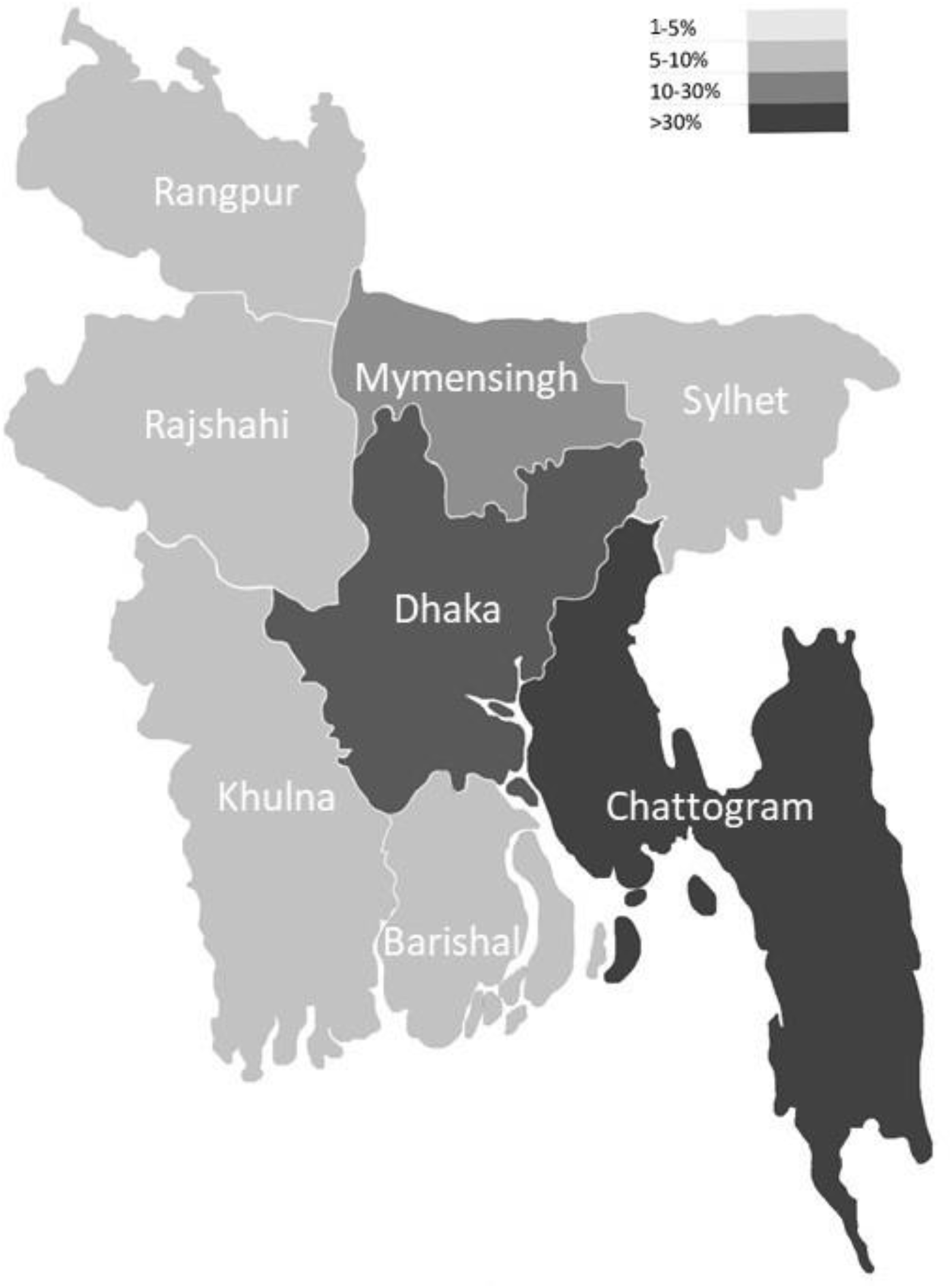
Heatmap showing the administrative division-wise distribution of all the participants (n=2905)

**Table S2.** Responses to the attitude question A1 [“Do you think that this new coronavirus will finally be under control completely?”] by sociodemographic characteristics and source of information

|  |  | Online non-medical, n=1097 |  |  | Online medical, n=382 |  |  |
| --- | --- | --- | --- | --- | --- | --- | --- |
|  |  | Yes | No | Don't know | Yes | No | Don't know |
|  |  | n (%) | n (%) | n (%) | n (%) | n (%) | n (%) |
| Socio-demographic characteristics |  |  |  |  |  |  |  |
| Age (years) | ≥18 - <35 | 546 (60.6) | 94 (10.4) | 261 (29) | 196 (56.5) | 59 (17) | 92 (26.5) |
|  | ≥35 - <55 | 109 (63) | 23 (13.3) | 41 (23.7) | 14 (50) | 2 (7.1) | 12 (42.9) |
|  | ≥55 | 15 (65.2) | 2 (8.7) | 6 (26.1) | 4 (57.1) | 2 (28.6) | 1 (14.3) |
| Gender | Male | 472 (63.5) | 86 (11.6) | 185 (24.9) | 103 (64) | 32 (19.9) | 26 (16.1) |
|  | Female | 198 (55.9) | 33 (9.3) | 123 (34.7) | 111 (50.2) | 31 (14) | 79 (35.7) |
| Education | ≤ Grade 5 | 0 (0) | 0 (0) | 2 (100) |  |  |  |
|  | Grade 6 to 12 | 36 (55.4) | 5 (7.7) | 24 (36.9) |  |  |  |
|  | > Grade 12 to Bachelor | 347 (60.8) | 66 (11.6) | 158 (27.7) | 157 (58.4) | 41 (15.2) | 71 (26.4) |
|  | > Bachelor to Master or above | 287 (62.5) | 48 (10.5) | 124 (27) | 57 (50.4) | 22 (19.5) | 34 (30.1) |
| Lived mostly in last 4 months | Village | 79 (62.2) | 12 (9.4) | 36 (28.3) | 21 (55.3) | 8 (21.1) | 9 (23.7) |
|  | Urban/town | 578 (60.8) | 105 (11.1) | 267 (28.1) | 191 (56.2) | 55 (16.2) | 94 (27.6) |
|  | Other | 13 (65) | 2 (10) | 5 (25) | 2 (50) | 0 (0) | 2 (50) |
| Financial condition | Poor | 25 (55.6) | 2 (4.4) | 18 (40) |  |  |  |
|  | Middle-class | 623 (61.6) | 110 (10.9) | 279 (27.6) | 197 (56.1) | 56 (16) | 98 (27.9) |
|  | Rich | 22 (55) | 7 (17.5) | 11 (27.5) | 17 (54.8) | 7 (22.6) | 7 (22.6) |
| Source of information |  |  |  |  |  |  |  |
| Television | Yes | 347 (61.4) | 58 (10.3) | 160 (28.3) | 104 (56.8) | 32 (17.5) | 47 (25.7) |
|  | No | 323 (60.7) | 61 (11.5) | 148 (27.8) | 110 (55.3) | 31 (15.6) | 58 (29.1) |
| Newspaper | Yes | 213 (63.8) | 39 (11.7) | 82 (24.6) | 68 (56.7) | 20 (16.7) | 32 (26.7) |
|  | No | 457 (59.9) | 80 (10.5) | 226 (29.6) | 146 (55.7) | 43 (16.4) | 73 (27.9) |
| Internet | Yes | 565 (59.8) | 112 (11.9) | 268 (28.4) | 189 (56.1) | 54 (16) | 94 (27.9) |
|  | No | 105 (69.1) | 7 (4.6) | 40 (26.3) | 25 (55.6) | 9 (20) | 11 (24.4) |
| Other people | Yes | 99 (60.7) | 22 (13.5) | 42 (25.8) | 45 (63.4) | 8 (11.3) | 18 (25.4) |
|  | No | 571 (61.1) | 97 (10.4) | 266 (28.5) | 169 (54.3) | 55 (17.7) | 87 (28) |
| Other ways | Yes | 28 (58.3) | 6 (12.5) | 14 (29.2) | 22 (52.4) | 5 (11.9) | 15 (35.7) |
|  | No | 642 (61.2) | 113 (10.8) | 294 (28) | 192 (56.5) | 58 (17.1) | 90 (26.5) |
| Total |  | 670 (61.1) | 119 (10.8) | 308 (28.1) | 214 (56) | 63 (16.5) | 105 (27.5) |

**Table S3.** Responses to the attitude question A2 [“This virus is created by humans” - this kind of discourse is heard. Do you believe that it is true?) by sociodemographic characteristics and source of information

|  |  | Online non-medical, n=1097 |  |  | Online medical, n=382 |  |  |
| --- | --- | --- | --- | --- | --- | --- | --- |
|  |  | Yes | No | Don't know | Yes | No | Don't know |
|  |  | n (%) | n (%) | n (%) | n (%) | n (%) | n (%) |
| Socio-demographic characteristics |  |  |  |  |  |  |  |
| Age (years) | ≥18 - <35 | 167 (18.5) | 231 (25.6) | 503 (55.8) | 64 (18.4) | 123 (35.4) | 160 (46.1) |
|  | ≥35 - <55 | 38 (22) | 45 (26) | 90 (52) | 5 (17.9) | 12 (42.9) | 11 (39.3) |
|  | ≥55 | 5 (21.7) | 4 (17.4) | 14 (60.9) | 0 (0) | 3 (42.9) | 4 (57.1) |
| Gender | Male | 146 (19.7) | 194 (26.1) | 403 (54.2) | 26 (16.1) | 68 (42.2) | 67 (41.6) |
|  | Female | 64 (18.1) | 86 (24.3) | 204 (57.6) | 43 (19.5) | 70 (31.7) | 108 (48.9) |
| Education | ≤ Grade 5 | 0 (0) | 0 (0) | 2 (100) |  |  |  |
|  | Grade 6 to 12 | 17 (26.2) | 15 (23.1) | 33 (50.8) |  |  |  |
|  | > Grade 12 to Bachelor | 112 (19.6) | 143 (25) | 316 (55.3) | 50 (18.6) | 95 (35.3) | 124 (46.1) |
|  | > Bachelor to Master or above | 81 (17.6) | 122 (26.6) | 256 (55.8) | 19 (16.8) | 43 (38.1) | 51 (45.1) |
| Lived mostly in last 4 months | Village | 23 (18.1) | 32 (25.2) | 72 (56.7) | 13 (34.2) | 14 (36.8) | 11 (28.9) |
|  | Urban/town | 184 (19.4) | 239 (25.2) | 527 (55.5) | 55 (16.2) | 122 (35.9) | 163 (47.9) |
|  | Other | 3 (15) | 9 (45) | 8 (40) | 1 (25) | 2 (50) | 1 (25) |
| Financial condition | Poor | 11 (24.4) | 14 (31.1) | 20 (44.4) |  |  |  |
|  | Middle-class | 191 (18.9) | 258 (25.5) | 563 (55.6) | 59 (16.8) | 130 (37) | 162 (46.2) |
|  | Rich | 8 (20) | 8 (20) | 24 (60) | 10 (32.3) | 8 (25.8) | 13 (41.9) |
| Source of information |  |  |  |  |  |  |  |
| Television | Yes | 103 (18.2) | 136 (24.1) | 326 (57.7) | 36 (19.7) | 62 (33.9) | 85 (46.4) |
|  | No | 107 (20.1) | 144 (27.1) | 281 (52.8) | 33 (16.6) | 76 (38.2) | 90 (45.2) |
| Newspaper | Yes | 60 (18) | 78 (23.4) | 196 (58.7) | 24 (20) | 38 (31.7) | 58 (48.3) |
|  | No | 150 (19.7) | 202 (26.5) | 411 (53.9) | 45 (17.2) | 100 (38.2) | 117 (44.7) |
| Internet | Yes | 178 (18.8) | 240 (25.4) | 527 (55.8) | 63 (18.7) | 116 (34.4) | 158 (46.9) |
|  | No | 32 (21.1) | 40 (26.3) | 80 (52.6) | 6 (13.3) | 22 (48.9) | 17 (37.8) |
| Other people | Yes | 22 (13.5) | 41 (25.2) | 100 (61.3) | 13 (18.3) | 27 (38) | 31 (43.7) |
|  | No | 188 (20.1) | 239 (25.6) | 507 (54.3) | 56 (18) | 111 (35.7) | 144 (46.3) |
| Other ways | Yes | 8 (16.7) | 11 (22.9) | 29 (60.4) | 7 (16.7) | 16 (38.1) | 19 (45.2) |
|  | No | 202 (19.3) | 269 (25.6) | 578 (55.1) | 62 (18.2) | 122 (35.9) | 156 (45.9) |
| Total |  | 210 (19.1) | 280 (25.5) | 607 (55.3) | 69 (18.1) | 138 (36.1) | 175 (45.8) |

